# The impact of asymptomatic COVID-19 infections on future pandemic waves

**DOI:** 10.1101/2020.06.22.20137489

**Authors:** Spencer J Fox, Remy Pasco, Mauricio Tec, Zhanwei Du, Michael Lachmann, James Scott, Lauren Ancel Meyers

**Affiliations:** The University of Texas at Austin, Austin, Texas 78712, The United States of America; Santa Fe Institute, Santa Fe, New Mexico, The United States of America

## Abstract

The prevalence of asymptomatic COVID-19 infections is largely unknown and may determine the course of future pandemic waves and the effectiveness of interventions. Using an epidemiological model fit to COVID-19 hospitalization counts from New York City, New York and Austin, Texas, we found that the *undocumented* attack rate in the first pandemic wave depends on the proportion of asymptomatic infections but not on the infectiousness of such individuals. Based on a recent report that 22.7% of New Yorkers are seropositive for SARS-CoV-2, we estimate that 56% (95% CI: 53-59%) of COVID-19 infections are asymptomatic. Given uncertainty in the case hospitalization rate, however, the asymptomatic proportion could be as low as 20% or as high as 80%. We find that at most 1.26% of the Austin population was infected by April 27, 2020 and conclude that immunity from undetected infections is unlikely to slow future pandemic spread in most US cities in the summer of 2020.

---

As of June 20, 2020, the COVID-19 pandemic has caused nearly 9 million reported infections and over 450,000 confirmed deaths ^1^. Statistical and serological estimates suggest the true prevalence of the virus, SARS-CoV-2, may be anywhere from 5 to 50 times the reported case count ^2–6^. The failure to detect infections stems from both the limited availability of tests and potentially large numbers of asymptomatic or mild infections who may not have cause to seek testing.

Asymptomatic infections are cases that do not develop clinical symptoms (e.g., fever or cough) but would likely test positive if given a SARS-CoV-2 nucleic acid test ^7–11^. Recent studies imply that anywhere from 5% to 95% of infections are mild or asymptomatic ^7,10,12–18^ and are equivocal about the role of such infections in community transmission ^10,12,16,19–22^. One study estimated that asymptomatic infections are 67% as infectious as symptomatic infections, but reported a confidence interval ranging from 29% to 142% ^12^. Uncertainty in estimates may stem from demographic differences in study populations–for example, younger populations may have higher asymptomatic rates ^7,10^, differences in classification of mild infections ^10,21^, or variability in the sensitivity and specificity of diagnostic tests ^8^.

Silent spread of COVID-19 has two key implications for managing future pandemic risks. First, asymptomatic transmission can amplify emerging clusters prior to their detection, particularly in younger populations ^23,24^. This poses a particular risk as schools and universities come back to campus in the fall of 2020. Second, high levels of undocumented cases may imply that cities are closer to achieving herd immunity than suggested by confirmed case counts ^25–27^.

To provide clarity on both of these issues–the extent of silent spread and implications for herd immunity–we fit a stochastic Susceptible-Exposed-Symptomatic/Asymptomatic-Recovered compartmental model to COVID-19 hospitalization data from Austin, Texas and New York City through April 27, 2020. We varied three key inputs–the asymptomatic proportion (from 0% to 99%), the relative infectiousness of asymptomatic infections (from 29% to 142%), and the symptomatic infection hospitalization rate (from 4.43% to 9.05% in Austin and 3.05% to 10.5% in NYC)–to assess the role and implications of asymptomatic SARS-CoV-2 infections (Supplement).

Across all inputs, we achieve good fits with hospital admission data and estimate that the reproduction number dropped dramatically between early March and April 27, by roughly 98-100% in NYC and 80-93% in Austin (Figure 1A, Figure S1). When we average the *R*_*t*_ values from the initial epidemic period up to the first hospitalization date, we find *R*_*t*_ values of 4.85 (95% CI: 2.04-12.0) for NYC and 5.87 (95% CI: 3.36-10.7) for Austin. However, these estimates depend on the case hospitalization rate, proportions of infections that are asymptomatic, and only slightly on the infectiousness of asymptomatic infections (Figure S2-S4).

**Figure 1:**
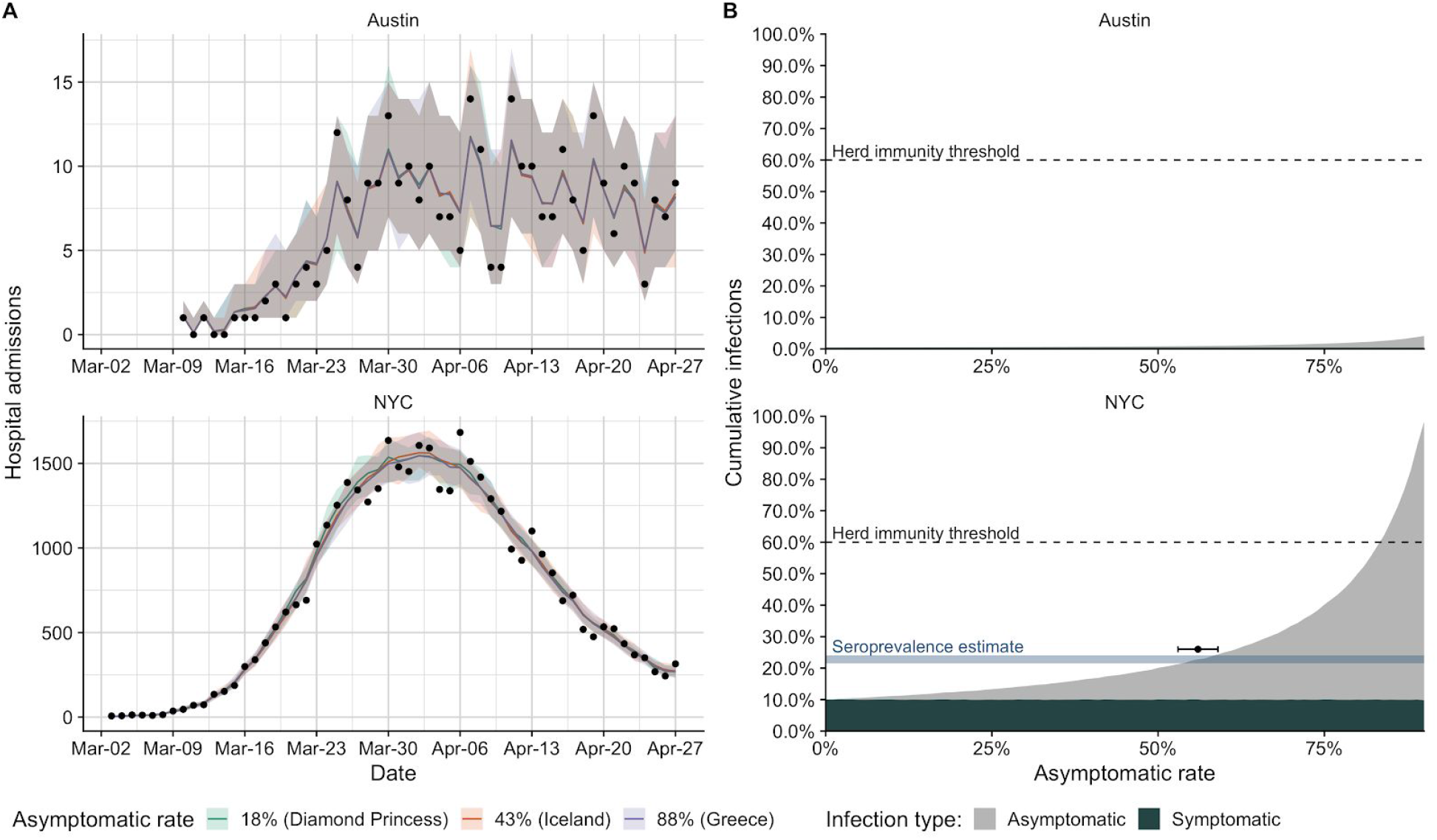
Projected COVID-19 hospitalizations and infections in Austin, TX and New York City, NY (NYC) based on hospitalization admissions through April 27, 2020. (A) Observed hospital admissions (points) versus estimated hospital admissions (lines) based on data-driven stochastic SEIR compartmental models of COVID-19 transmission in Austin (top) and NYC (bottom). The colored lines and shading indicate mean estimates and 95% prediction intervals assuming three different asymptomatic proportions ^33–35^. Corresponding estimates for the reproduction number (*R*_*t*_) and cumulative infections are provided Figure S1. (B) Proportion of population infected by SARS-CoV-2 in Austin (top) and NYC (bottom) through April 27, 2020, stratified by symptomatic (dark slate) versus asymptomatic (grey). Simulations assume a symptomatic hospitalization rate of 4.43% for Austin and 5.13% for NYC, as estimated in the supplemental information ^29^. Across all asymptomatic rates, Austin is expected to have an exceedingly low proportion infected. Horizontal dashed lines indicate the approximate herd immunity threshold above which the pandemic is expected to subside ^32^. In NYC, an asymptomatic rate above 83% might imply that the city may be close to achieving herd immunity. The horizontal blue bar indicates a serology-based estimate for the cumulative incidence in NYC by April 29th (21.5%-24%) ^28^. The horizontal black crossbar indicates where this estimate intersects with our projections (top of the gray curve) and suggests an asymptomatic rate in NYC of 56% (53%-59%) as described in the supplemental information.

To triangulate the asymptomatic proportion, we consider a recent serology-based estimate that 22.7% (95% CI: 21.5-24.0%) of the NYC population was infected as of April 28, 2020 ^28^. Assuming that 5.1% of symptomatic infections in NYC were hospitalized ^29^, we estimate the asymptomatic proportion to be 56% (53-59%) (Figure 1B). However, the range of plausible values widens to 20%-80% when we account for the uncertainty in the symptomatic hospitalization rate (Figure S6).

Assuming asymptomatic COVID-19 infections are fully immunizing ^30,31^, previous studies suggest that at least 60% of the population will need to be infected to reach herd immunity–the point where enough people have been immunized that the pandemic stops spreading ^32^. Our initial estimates for early transmission rates in New York City and Austin suggest a higher potential value of 82% 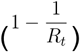, but we use 60% as a conservative estimate. The key question is whether asymptomatic infections have already begun to close the gap between the small numbers of reported cases and the herd immunity threshold. In Austin, the answer is clearly *no*. Our estimates suggest it’s only possible in our most extreme scenario where 99% of infections are asymptomatic and the symptomatic hospitalization rate is 2.6% (Figure S5). Based on the asymptomatic proportion derived from the NYC serology data, we estimate that 0.94% (95% CI: 0.72%-1.26%) of the Austin population was infected by April 27, 2020. These estimates are consistent across symptomatic hospitalization rates and imply a 9.7% (95% CI: 7.2-12.3%) infection detection rate in Austin. Interestingly, estimates of current immunity levels do not depend on the relative infectiousness of asymptomatic infections (Figure S6).

The proportion of SARS-CoV-2 infections that are asymptomatic remains uncertain, but we have narrowed the plausible range of values from 5-95% ^7,10,12–16^ to 20-80%, with a most likely value of 56%. Moreover, our analysis provides four clear insights regarding the future spread of COVID-19 in US cities. First, in communities that slowed spread early and reported relatively small initial pandemic waves, like Austin, the vast majority of people remain susceptible to infection. If social distancing and other individual precautionary measures are fully relaxed, we would expect the reproduction number of the virus to rebound toward initial levels. Thus, such cities remain highly vulnerable to rapid transmission and hospital surges. Second, as case counts climb, the asymptomatic rate has an increasing impact on the fate of the epidemic. If we take our most likely estimate that 56% of infections are asymptomatic, then we expect cities to reach herd immunity after roughly 26.4% of the population have had symptomatic infections. Third, while estimates for total infections in Austin our insensitive to symptomatic hospitalization rate, this key parameter can modulate our estimates of the asymptomatic rate. Our estimates will become more precise as more more comprehensive and retrospective analyses of symptomatic infections become available. Finally, our estimates for total infections to date are remarkably insensitive to what we assume about the infectiousness of asymptomatic infections. However, understanding the role of asymptomatic infections in the transmission cycle will be critical to developing safe and sustainable reopening policies.

## Data Availability

All data and code available on request. Currently working on a clean github repository to store it.

## Author contributions

SJF, RP, ML, JS, LAM conceived and designed the study. RP, MT, ML, and JS provided guidance on computational work, and SJF carried out all analyses. SJF wrote the first draft. LAM and JS supervised the work. All authors contributed to data interpretation and critical revision of the manuscript.

## Supplemental Information

### COVID-19 stochastic transmission model

We model the transmission dynamics of COVID-19 using a stochastic SEIR compartmental model that explicitly accounts for symptomatic and asymptomatic infectious compartments, hospitalizations, and deaths. The model is defined by the following set of equations.

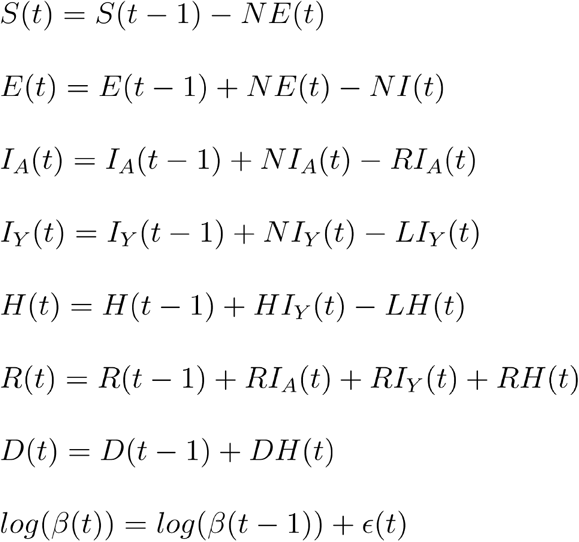

and

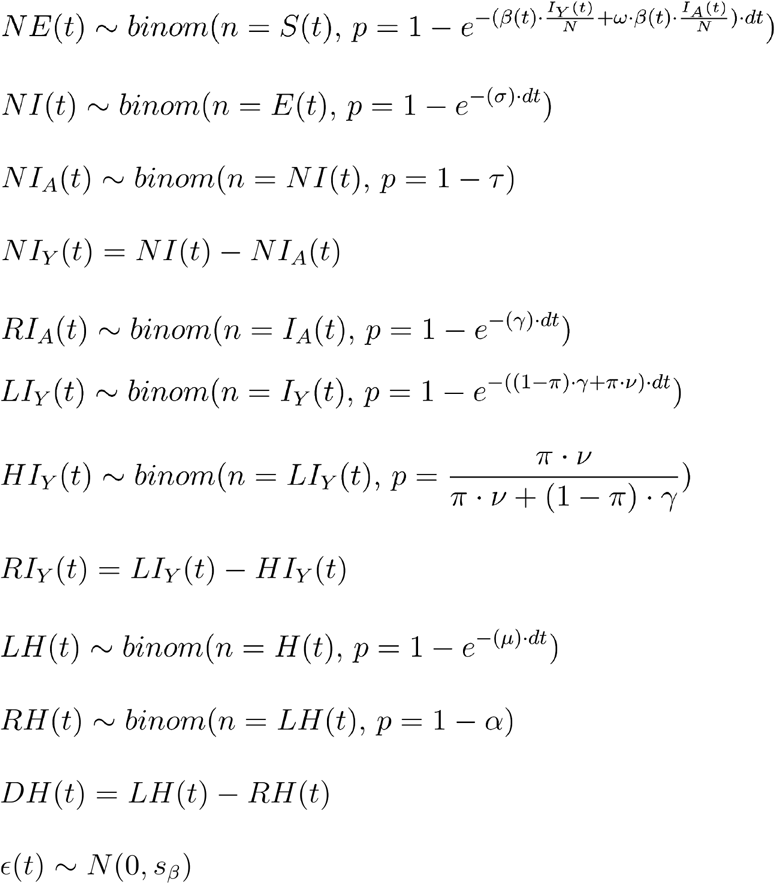

Where 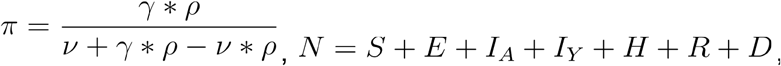, and with parameters asdefined in the parameter table (Table S1) and states as defined in the state table (Table S2).

**Table S1:** Model parameter definitions and estimates

| Parameter | Definition | Estimate | Citation |
| --- | --- | --- | --- |
| $\beta(t)$ | Daily transmission rate | Estimated from data | |
| $s_\beta$ | Standard deviation controlling the transmission rate AR(1) process on a log scale | Estimated from data | |
| $t_0$ | Epidemic start date | Fixed at estimated value:<br>Austin = February 18, 2020<br>NYC = January 22, 2020 | |
| $t_d$ | Duration of the simulation in days<br>(April 27, 2020 - $t_0$) | Austin = 72<br>NYC = 98 | |
| $\omega$ | Relative infectiousness of asymptomatic individuals compared with symptomatic individuals. | (0.29, 0.67, 1.42) | 12 |
| $\tau$ | Proportion of cases that are symptomatic | (0.01, 0.02, ..., 0.99, 1.0) | |
| $\sigma$ | Rate of becoming infectious following exposure to COVID-19 | $\frac{1}{2.9 \text{ days}}$ | 36 |
| $\gamma$ | Recovery rate while infectious | $\frac{1}{7 \text{ days}}$ | 36 |
| $\rho$ | Proportion of symptomatic cases that require hospitalization normalized to city demographic data | Austin = 2.64%, 4.43%, 9.05%<br>NYC = 3.05%, 5.13%, 10.5% | 29,37 |
| $\nu$ | Rate of hospitalization of symptomatic infections | $\frac{1}{7 \text{ days}}$ | 38 |
| $\alpha$ | Proportion of hospitalizations that die | Austin = 0.154<br>NYC = 0.21 | 39,40 |
| $\mu$ | Rate of leaving hospital | Austin = $\frac{1}{10.5 \text{ days}}$<br>NYC = $\frac{1}{4.1 \text{ days}}$ | 39,40 |
| $R(t)$ | Effective reproduction number at time $t$ | $(\frac{\tau \cdot \beta(t)}{(1-\pi) \cdot \gamma + \pi \cdot \nu} + \frac{(1-\tau) \cdot \beta(t) \cdot \omega}{\gamma}) \cdot \frac{S(t)}{N}$ | |

**Table S2:** Model state definitions and initial conditions

| State | Definition | Initial value |
| --- | --- | --- |
| $S$ | Individuals susceptible to COVID-19 | Austin = 2,168,316<br>NYC = 8,398,747 |
| $E$ | Individuals exposed to COVID-19 but not currently infectious | Austin = 0<br>NYC = 0 |
| $I_A$ | Infectious individuals who are asymptomatic | Austin = 0<br>NYC = 0 |
| $I_Y$ | Infectious individuals who are symptomatic | Austin = 1<br>NYC = 1 |
| $H$ | Hospitalized individuals | Austin = 0<br>NYC = 0 |
| $R$ | Recovered individuals | Austin = 0<br>NYC = 0 |
| $D$ | Individuals who have died | Austin = 0<br>NYC = 0 |

### Calculating the symptomatic hospitalization rate

We used estimates of the expected hospitalization rate by age for infected cases from early data from China as a proxy for the estimated symptomatic infection hospitalization rate ^29^. Using the age distribution from Austin and NYC (Table S3), we calculated the expected symptomatic hospitalization rate for each city by multiplying the estimated hospitalization rate for each age group by the fraction of the population in the age group and taking the sum of all age groups. Population distribution in the cities was estimated using 2015 census data as made accessible through the tidycensus package in R ^41,42^.

**Table S3:** Data for estimating the symptomatic hospitalization rate.

| Age group | Hospitalization rate (95% CI) <sup>29</sup> | Austin population | NYC population |
| --- | --- | --- | --- |
| 0-9 | 0% (0-0%) | 13.2% | 12.2% |
| 10-19 | 0.0408% (0.0243-0.0832%) | 13.2% | 10.9% |
| 20-29 | 1.04% (0.622-2.13%) | 15.8% | 16.1% |
| 30-39 | 3.43% (2.04-7%) | 16.7% | 15.7% |
| 40-49 | 4.25% (2.53-8.68%) | 14.2% | 12.9% |
| 50-59 | 8.16% (4.86-16.7%) | 11.7% | 12.6% |
| 60-69 | 11.8% (7.01-24%) | 8.67% | 10.1% |
| 70-79 | 16.6% (9.87-33.8%) | 4.33% | 5.82% |
| 80+ | 18.4% (11-37.6%) | 2.11% | 3.71% |

### Data for fitting

We obtained hospitalization data through personal communication from the City of Austin, which included confirmed daily COVID-19 admits and discharges from the hospitals in the MSA, and included data from Bastrop, Caldwell, Hays, Travis, Williamson counties in aggregate. NYC data were compiled by the Department of Health and Mental Hygiene (DOHMH) Incident Command System for COVID-19 Response (Surveillance and Epidemiology Branch in collaboration with Public Information Office Branch) and were obtained through the public repository from the NYC Department of health ^43^. The NYC data analyzed included daily hospital admit numbers for the city beginning on March 3rd.

### Epidemic start date

We could not estimate the epidemic start date directly using our model, because the transmission rate flexibility gave rise to similarly good fits within a wide-range of potential values for *t*_0_. We therefore conducted an independent estimation procedure to obtain reasonable epidemic start dates for both Austin and NYC. For each city we first obtained the number of hospital admits on the first day of data (Austin = 1, NYC = 6). We then used our best guess parameters for each city as described in Table S1 and chose *β* (0) = 0.67 as it produced 3 day doubling rate in cumulative cases and gave *R*_*e*_ *(0) = 4* which are consistent with observations across many locations for the early outbreak dynamics ^44^. We ran 1,000 stochastic simulations with these initial conditions, and identified the wait time for which the simulation first reached the number of admits in the first day of data for each city (1 admit for Austin and 6 admits for NYC). We estimated the start time from the resulting distribution of wait times for Austin as February 17, 2020 (IQR = February 11 - February 23) and for NYC as January 21, 2020 (IQR = January 12 - February 2).

### Model fitting procedure

We carried out two slightly different estimation procedures for NYC and Austin due to the differences in available data. For both cities we varied and fixed the symptomatic proportion (*τ*), the relative infectiousness of asymptomatic individuals (*ω*), and the symptomatic hospitalization rate (*ρ*). Ranges for these parameters were chosen based on literature estimates (Table S1).

### Austin likelihood

We obtained daily hospital admit (*H*_*A*_(*t*)) and discharge data (*H*_*D*_(*t*)), which included discharge due to both recovery and death, for the Austin MSA. In this model and for each combination of *ω, τ*, and *ρ* we estimated *β*(*t),S*_*β*_, and *μ* and fixed all parameters as described for Austin in Table S1. We assumed that both hospital admits and discharges were Poisson distributed around their predicted values from the stochastic model, and chose an informative, but relatively disperse prior for *S*_*β*_, to prevent the model from overfitting data through large transmission rate perturbations. The likelihood for the Austin model was thus:

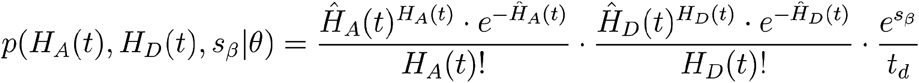

Where *Ĥ*_*A*_(*t*) and (*Ĥ*_*D*_(*t*)) are the predicted hospital admissions for day *t,t*_*d*_ is the duration of the epidemic, and θ refers to the fixed parameters from Table S1.

### NYC likelihood

We obtained daily hospital admit (*H*_*A*_(*t*)) data for NYC. Admit data for NYC was more overdispersed than that from Austin, with data at the peak fluctuating 20% or more on a daily basis. We therefore assumed that admit data were distributed according to a negative binomial distribution with mean *Ĥ*_*A*_(*t*) and dispersion parameter *r*. In this model and for each combination of *ω, τ* and *ρ* we estimated *β*(*t),S*_*β*_,*r* and fixed all parameters as described for NYC in Table S1. Similar to Austin, we chose an informative, but relatively disperse prior for *S*_*β*_, to prevent the model from overfitting data through large transmission rate perturbations. The likelihood for the NYC model was thus:

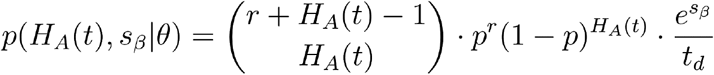

Where *r* is the estimated dispersion parameter, 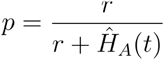, and *θ* refers to the fixed parameters from Table S1.

### Fitting method

For each combination of parameters that were fixed by design, we fitted the remaining free parameters in the model using the iterated filtering algorithm made available through the mif2 function in the pomp package in R ^45,46^. This algorithm is a stochastic optimization procedure; it performs maximum likelihood estimation using a particle filter to provide a noisy estimate of the likelihood for a given combination of the parameters. For each parameter combination we ran 1,000 iterations of iterated filtering, each with 10,000 particles. We repeated this fitting process five times for each combination of fixed parameters. As the stochastic fitting procedure can sometimes stick in local maxima or impossible parameter combinations, we selected the fitted model with the highest likelihood out of the five independent runs as our best fit model. Then fixing the best fit maximum likelihood parameter estimates, we calculated smoothed posterior estimates for all of the states within the model through time (including *β* (*t*) which is technically a state variable in our model formulation, as it changes through time according to a stochastic process). We calculated these smoothed posteriors as follows:

1. We ran 1,000 independent particle filters at the MLE, each with 10,000 particles. For each run, *k*, of particle filtering, we kept track of the complete trajectory of each particle, as well as the filtered estimate of the likelihood,*L*_*k*_.
2. For each of the 1,000 particle filtering runs, we randomly sampled a single complete particle trajectory, giving us 1,000 separate trajectories for all state variables.
3. We resampled from these trajectories with probabilities proportional to *L*_*k*_ to give a distribution of state trajectories

The result can be thought of as an empirical-Bayes posterior distribution: that is, a set of 1,000 smoothed posterior draws from all state variables, conditional on the maximum likelihood estimates for the model’s free parameters. This smoothed posterior distribution is how we calculate means and credible intervals for *β* (*t*) in addition to all other time-varying state variables. In general our estimates were more stable for Austin, potentially due to the high accuracy in the underlying data, or because we were able to fit both hospital admits and discharges.

### Estimating the asymptomatic rate

For each symptomatic hospitalization rate we calculated the mean and 95% confidence interval by comparing our fitted model estimates with the seroprevalence estimates from NYC. For a given hospitalization rate, we fitted 99 models assuming 0-99% asymptomatic rates for NYC as described above. We then calculated the mean and 95% confidence interval for the final estimate of the total infected population. Our mean estimate for the asymptomatic rate was then the asymptomatic rate whose mean value was most similar to the NYC estimate of 22.7% prevalence. 95% confidence intervals were calculated as the minimum and maximum asymptomatic rate whose fitted models overlapped with the seroprevalence confidence interval of 21.5% to 24.0%. Point estimates and confidence intervals for each asymptomatic rate can be seen in Figure S5, while asymptomatic rate estimates can be seen in Figure S6.

---

**Figure S1:**
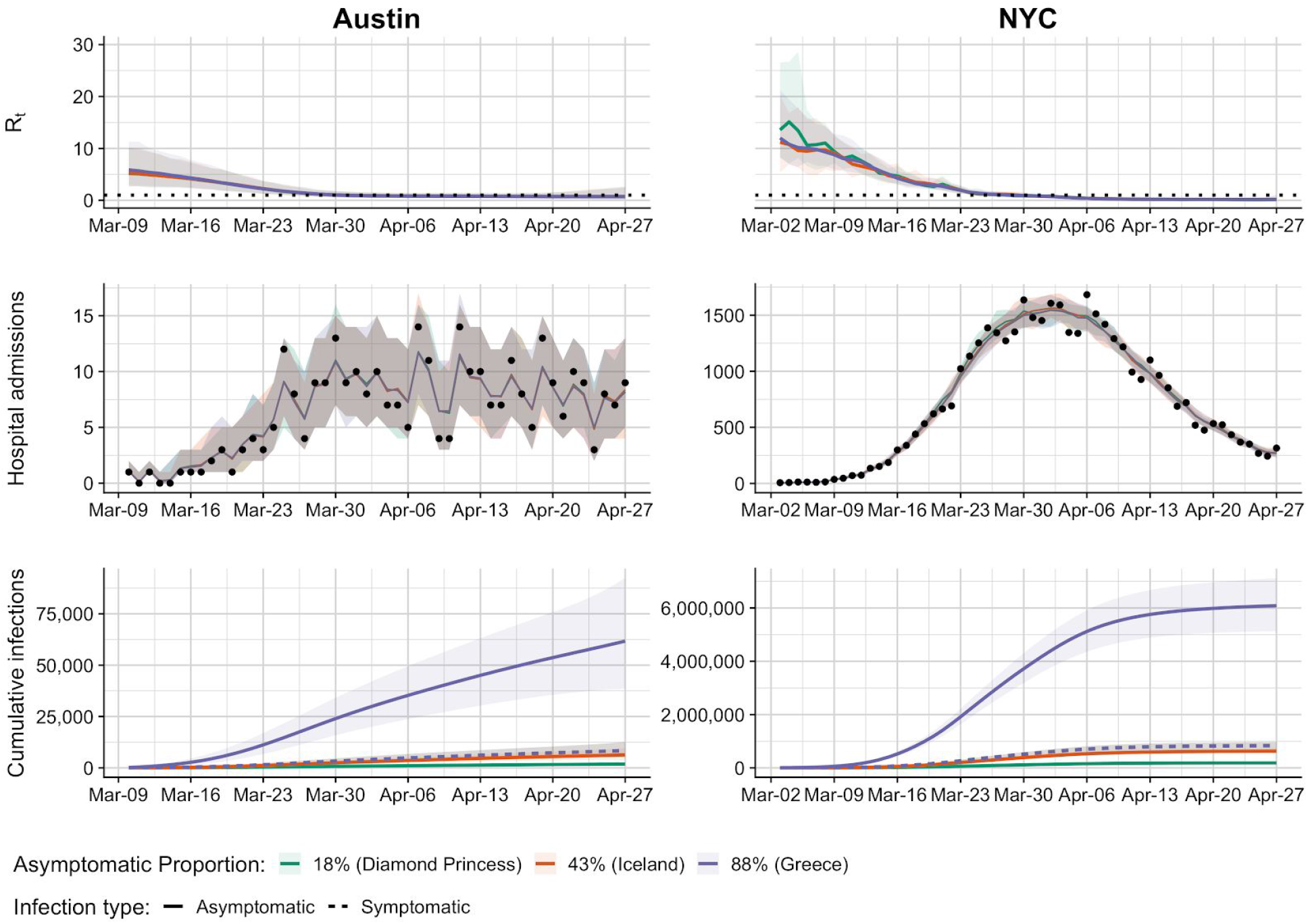
Fitted epidemic dynamics for Austin and NYC. *R*_*t*_ estimates (top), fitted hospital admit estimates (middle), and estimated cumulative incidence (bottom) for Austin (left) and NYC (right). *R*_*t*_ estimates are shown for a wide-range of estimated asymptomatic proportions (colors) from the recent literature ^33–35^. Hospital admission data (points) are compared with posterior distributions from the best fit model. Cumulative infections are shown for both asymptomatic (solid line) and symptomatic infections (dashed line), with symptomatic infection estimates overlapping across the range of asymptomatic rates.

**Figure S2:**
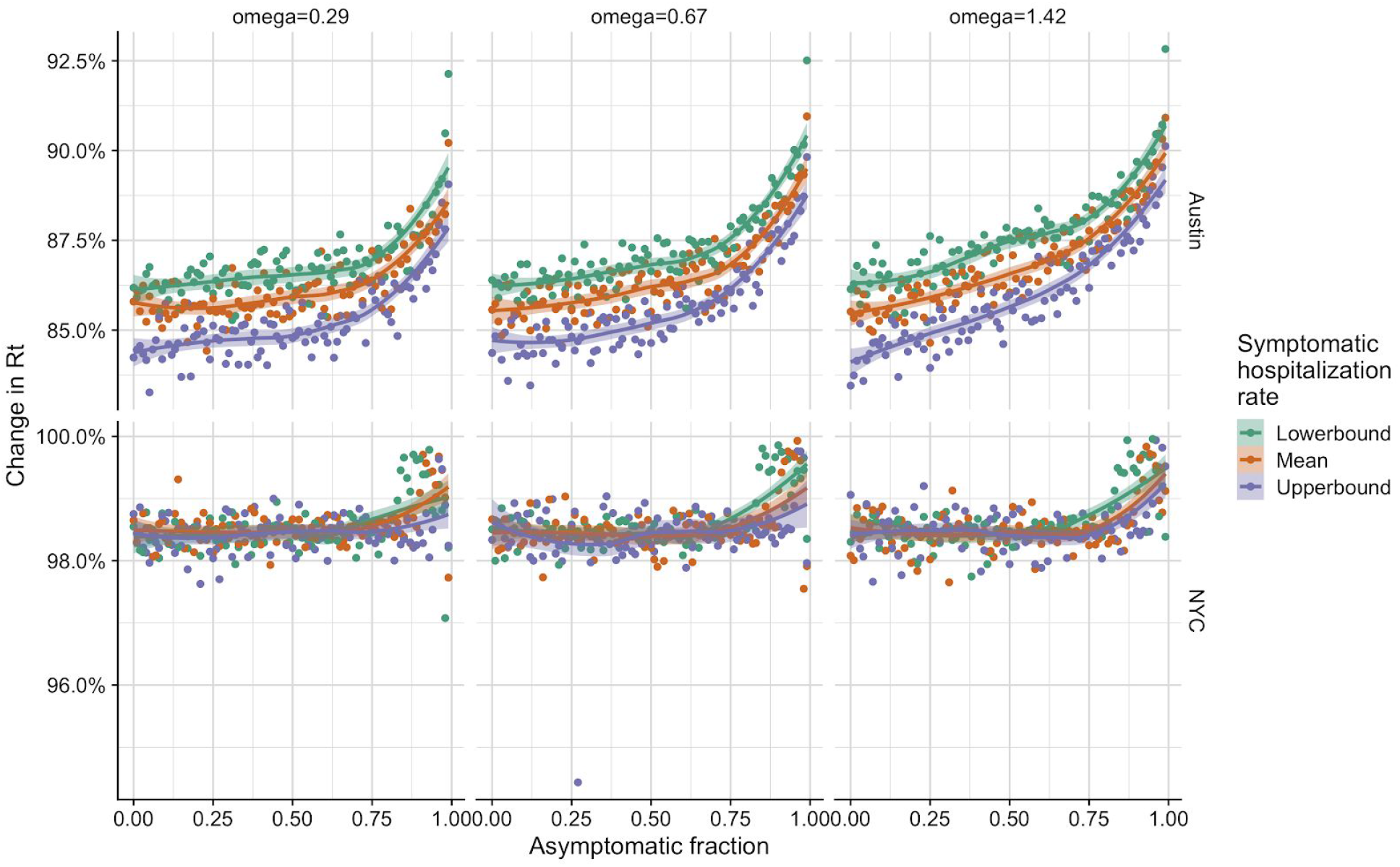
Reduction in R_t_ across the epidemics for Austin and NYC. Each panel corresponds with a specific city (row) and relative infectiousness of asymptomatic individuals (column-*ω*). Points correspond with estimated drop in *R*_*t*_ between the first and last dates of hospitalization data for each city. Colors correspond with estimated rate of hospitalization for symptomatic individuals, and smoothed splines are included to identify trends. In general Austin estimates are slightly more stable than those from NYC, likely do to the higher quality and availability of data. Symptomatic hospitalization rates vary between Austin and NYC due to different demographic makeups and the values are estimated to be 2.64%, 4.43%, and 9.05% for Austin and 3.05%, 5.13%, and 10.5% for NYC for lowerbound, mean, and upperbound respectively.

**Figure S3:**
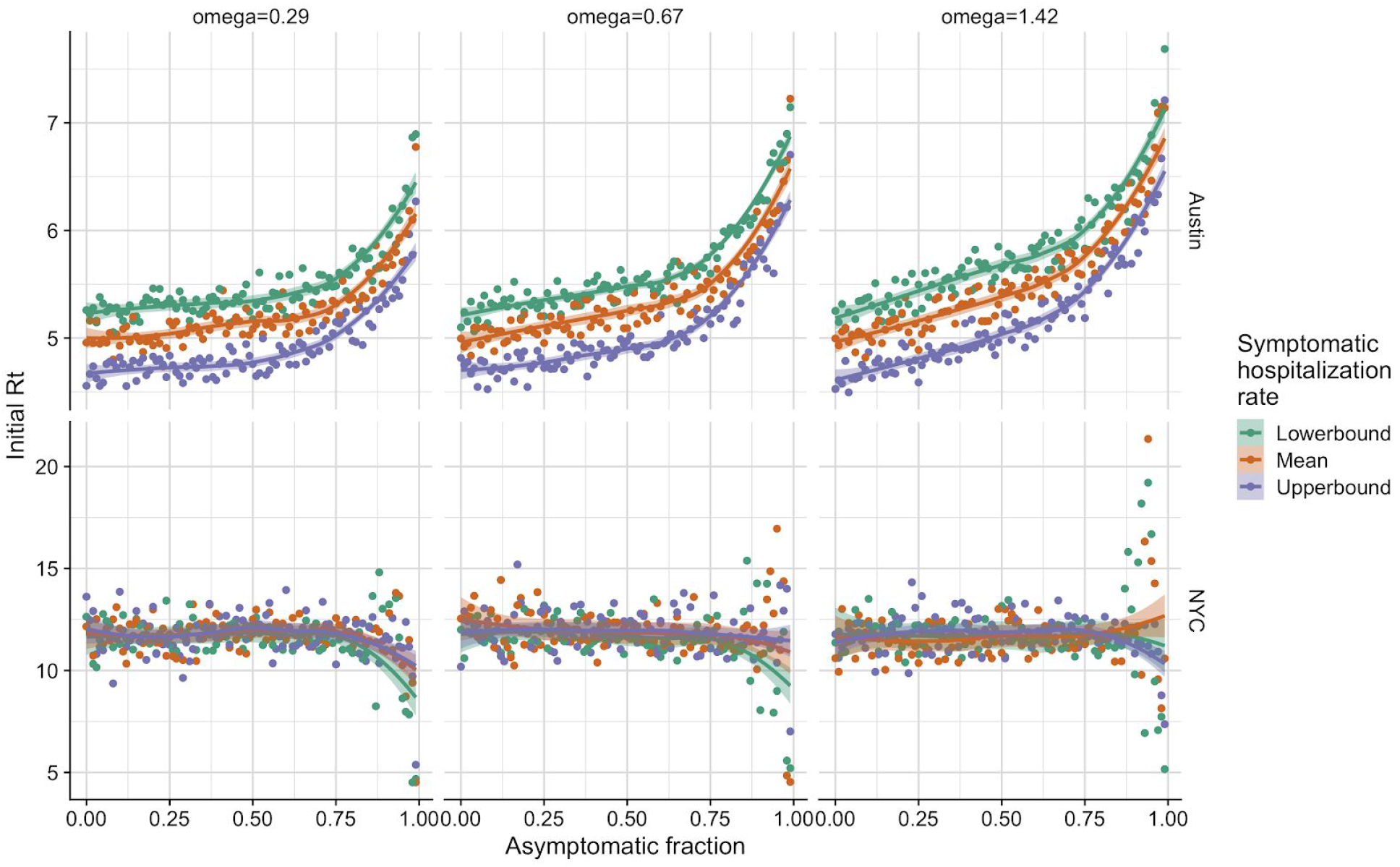
Initial estimates for R_t_ across the epidemics for Austin and NYC. Each panel corresponds with a specific city (row) and relative infectiousness of asymptomatic individuals (column-*ω*). Points correspond with the initial estimated *R*_*t*_ on the first day of hospitalization data for each city (NYC = March 3, 2020, Austin = March 10, 2020). Colors correspond with estimated rate of hospitalization for symptomatic individuals, and smoothed splines are included to identify trends. In general Austin estimates are slightly more stable than those from NYC, likely do to the higher quality and availability of data. Symptomatic hospitalization rates vary between Austin and NYC due to different demographic makeups and the values are estimated to be 2.64%, 4.43%, and 9.05% for Austin and 3.05%, 5.13%, and 10.5% for NYC for lowerbound, mean, and upperbound respectively.

**Figure S4:**
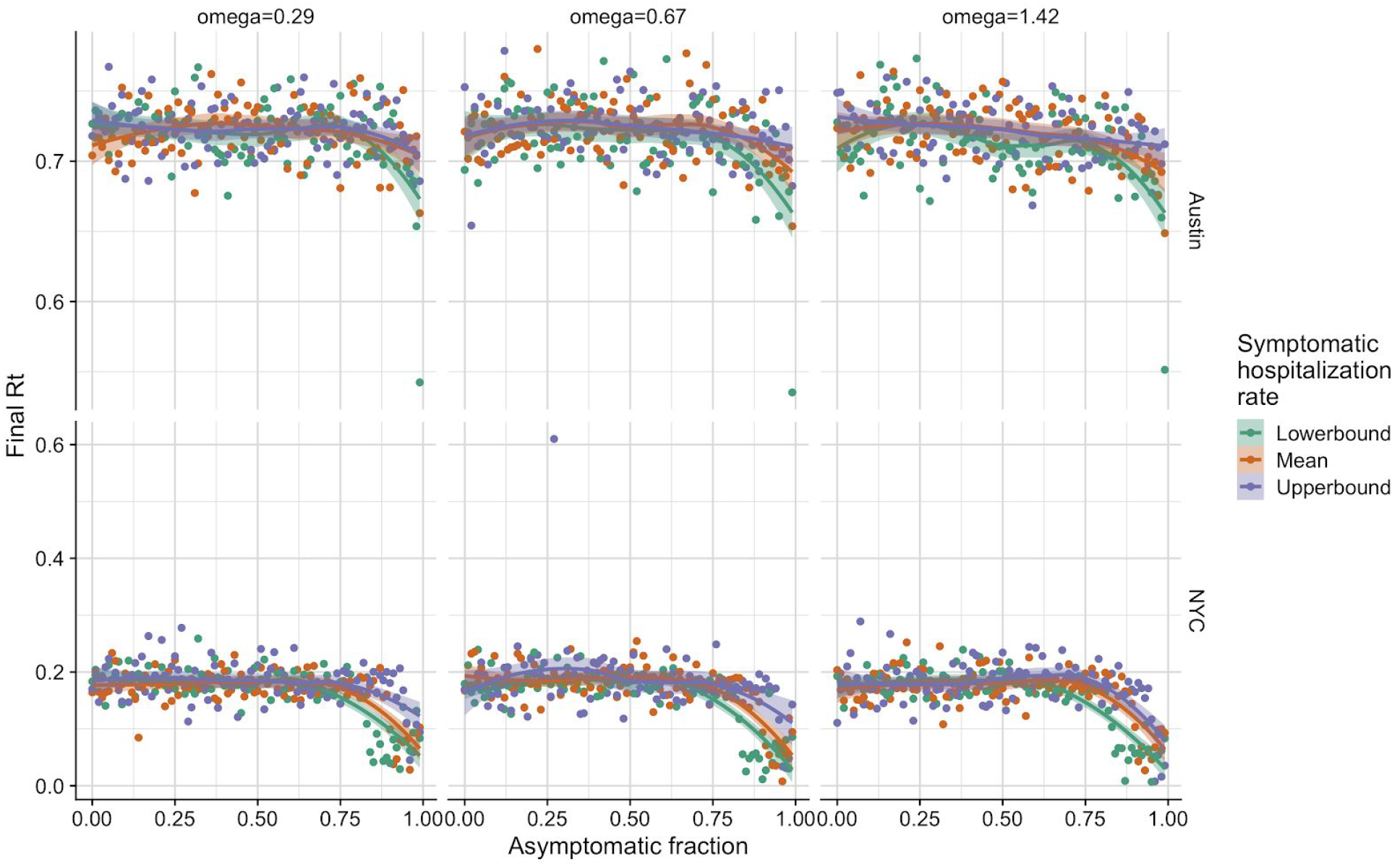
Final estimates for R_t_ across the epidemics for Austin and NYC. Each panel corresponds with a specific city (row) and relative infectiousness of asymptomatic individuals (column-*ω*). Points correspond with estimated final *R*_*t*_ on April 27, 2020 for each city. Colors correspond with estimated rate of hospitalization for symptomatic individuals, and smoothed splines are included to identify trends. In general Austin estimates are slightly more stable than those from NYC, likely do to the higher quality and availability of data. Symptomatic hospitalization rates vary between Austin and NYC due to different demographic makeups and the values are estimated to be 2.64%, 4.43%, and 9.05% for Austin and 3.05%, 5.13%, and 10.5% for NYC for lowerbound, mean, and upperbound respectively.

**Figure S5:**
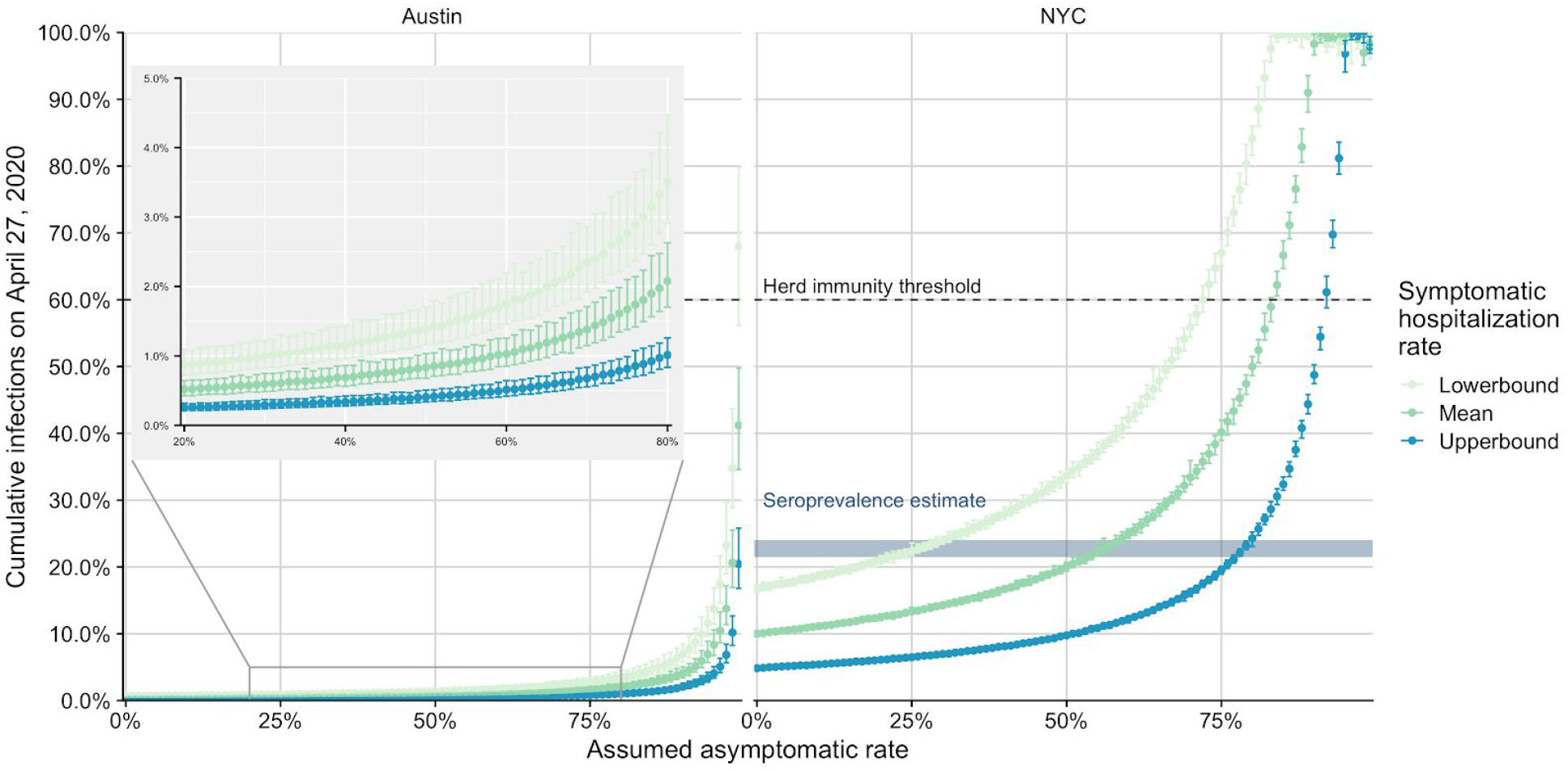
Estimated cumulative infections for Austin and NYC up to April 27, 2020 across asymptomatic rates and three different assumed symptomatic hospitalization rates (colors). Points and error bars represent the mean and 95% confidence interval for the estimated cumulative infections for the fitted model for Austin and NYC. Symptomatic hospitalization rates vary between Austin and NYC due to different demographic makeups and the values are estimated to be 2.64%, 4.43%, and 9.05% for Austin and 3.05%, 5.13%, and 10.5% for NYC for lowerbound, mean, and upperbound respectively. Estimates derived from Asymptomatic rates range from 0-99%. Horizontal dashed line indicates an estimate for herd immunity which is reached when cumulative infections are above the line, assuming fully immunizing infections and *R*_*0*_=2.5. Horizontal rectangle in NYC plot corresponds with estimated cumulative incidence on April 29th, 2020 ^28^. For each hospitalization rate, we estimate the asymptomatic rate that matches the seroprevalence data by identifying the range of asymptomatic rates whose confidence interval spans the confidence interval from the seroprevalence study. Inset plot zooms in for Austin data that spans the uncertainty in asymptomatic rates estimated from NYC (20-80% asymptomatic). These estimates assume asymptomatic infectiousness of 67% of that of symptomatic individuals, with other assumed infectiousness values not shown but mirroring the ones shown here.

**Figure S6:**
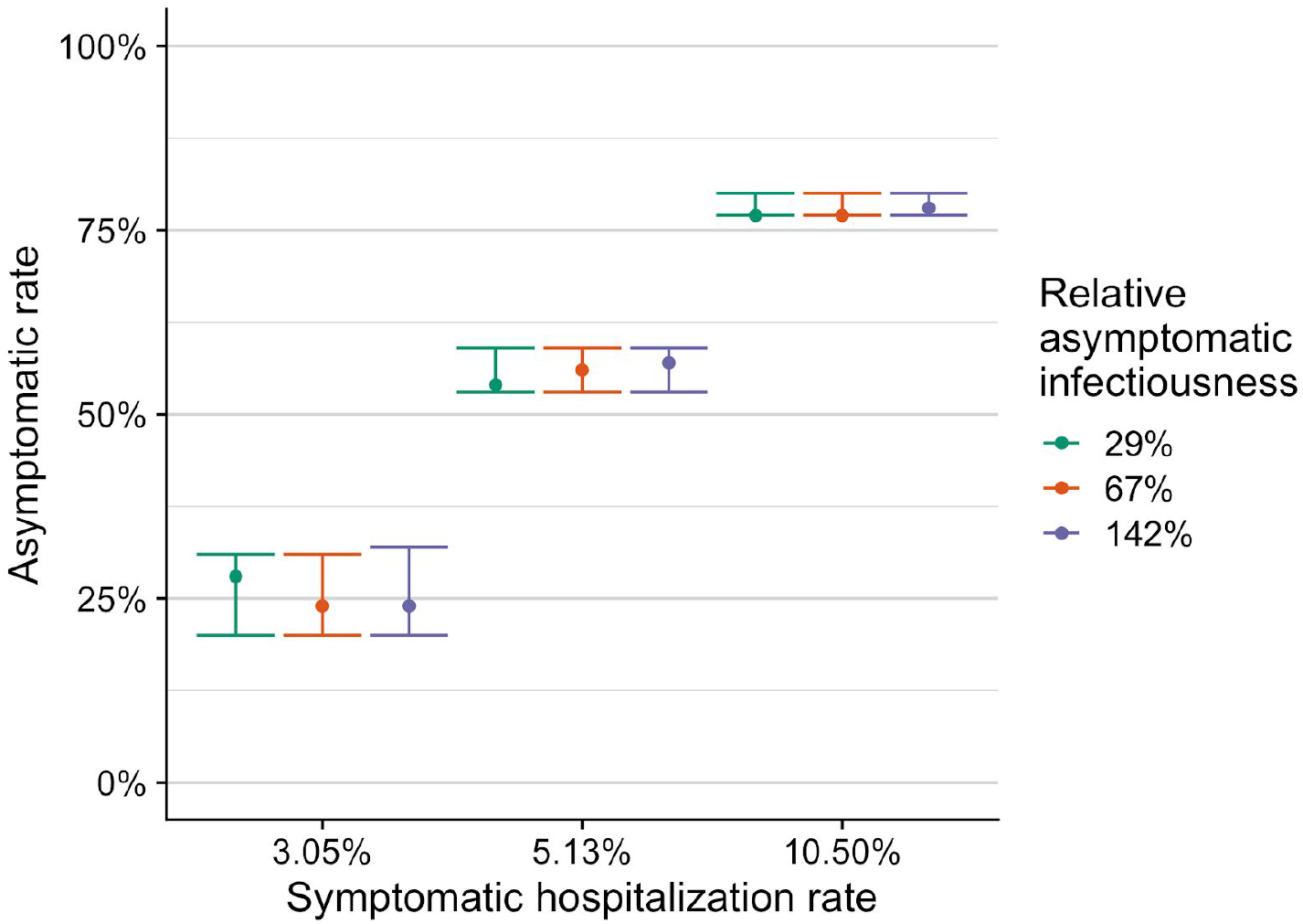
Asymptomatic rate estimate and 95% confidence interval from fitted model results across the mean and 95% confidence interval for symptomatic hospitalization rate. Colors indicate estimates made using different relative asymptomatic infectiousness estimates. Estimates obtained as described in the supplemental information.

